# Genomic characterization uncovers transmission dynamics of Marburg Virus in Rwanda following a single zoonotic spillover event

**DOI:** 10.1101/2024.11.01.24316374

**Authors:** Yvan Butera, Leon Mutesa, Edyth Parker, Raissa Muvunyi, Esperance Umumararungu, Alisen Ayitewala, Jean Pierre Musabyimana, Alhaji Olono, Placide Sesonga, Olusola Ogunsanya, Emmanuel Kabalisa, Oluwatobi Adedokun, Nelson Gahima, Laetitia Irankunda, Chantal Mutezemariya, Richard Niyonkuru, Arlene Uwituze, Ithiel Uwizera, James Kagame, Arlette Umugwaneza, John Rwabuhihi, Fidele Umwanankabandi, Valens Mbonitegeka, Edouard Ntagwabira, Etienne Kayigi, Gerard Izuwayo, Herve Murenzi, Therese Mukankwiro, Nasson Tuyiringire, Jean Marie Vianney Uwimana, Agnes Gasengayire, Reuben Sindayiheba, Glory-Ugochi Onyeugo, Merawi Aragaw, Lenny Gitundu, Radjabu Bigirimana, Mosoka Fallah, Adaora Ejikeme, Senga Sembuche, Alice Kabanda, Jean Claude Mugisha, Emmanuel Edwar Siddig Francis, Pierre Gashema, Jerome Ndayisenga, Alexis Rugamba, Faustin Kanyabwisha, Gad Murenzi, Anise Happi, Jean Claude Semuto Ngabonziza, Misbah Gashegu, Ayman Ahmed, Noella Bigirimana, Edson Rwagasore, Muhammed Semakula, Jean Paul Rwabihama, Clarisse Musanabaganwa, Eric Seruyange, Menelas Nkeshimana, Theogene Twagirumugabe, David Turatsinze, Eric Remera, Noel Gahamanyi, Sofonias Kifle Tessema, Isabelle Mukagatare, Sabin Nsanzimana, Christian Happi, Claude Mambo Muvunyi

## Abstract

The ongoing outbreak of Marburg virus disease (MVD) in Rwanda marks the third largest historically, though it has exhibited the lowest fatality rate. Genomic analysis has identified a lineage with limited internal diversity most closely related to a genome sequence from a sporadic case sampled in 2014 in Uganda, though the lineages have diverged from a common ancestor that was circulating for decades in the animal reservoir.

Notably, the data also provide evidence that the outbreak resulted from a single zoonotic transmission event with limited human-to-human transmission, rather than multiple independent zoonotic transmission events. The Rwandan MVD outbreak prompted a thorough investigation that included contact tracing, clinical assessment, travel history, sequencing, and serology testing, to trace the virus’s origin. Results of investigations linked the index case to a mining cave inhabited by *Rousettus aegyptiacus* (the Egyptian fruit bat), where three individuals tested seropositive for IgG and IgM, further supporting the zoonotic origin of the outbreak through human-animal interactions.

## Background

Since its discovery in 1967, the Marburg virus disease (MVD) has emerged as a significant global health threat, resulting in several outbreaks characterized by alarmingly high case fatality rates ranging from 22% to 90% (1). The first reported instances of MVD occurred in simultaneous outbreaks in Marburg and Frankfurt, Germany, as well as in Belgrade, Yugoslavia (now Serbia) (2). To date, there have been 19 recorded outbreaks, 563 confirmed cases, and 428 deaths worldwide (4). On September 27, 2024, Rwanda reported its first ever cases of MVD (5). The ongoing outbreak in Rwanda has confirmed 66 cases, with a case fatality rate of approximately 23% (5).

MVD has two distinct variants: Marburg virus (MARV) and Ravn virus (RAVV), with MARV being the more prevalent variant, responsible for the majority of sporadic MVD outbreaks worldwide. MARV is a negative-sense single-stranded RNA virus from the Filoviridae family, similar to Ebola and possesses inverse-complementary 3′ and 5′ termini (3). MVD is predominantly a zoonosis, transmitting to humans from either the bat natural reservoir or through intermediate hosts, including non-human primates. The mechanisms by which bats regulate viral replication and maintain complex immunity in contrast to humans remain unclear due to a lack of bat-specific research tools necessary for comparative studies, such as antibodies for flow cytometry, genomics, and transcriptomics (1,2). Studies confirm that MARV’s natural reservoir is the Egyptian fruit bat (*Rousettus aegyptiacus*) (6). The virus can cause viral hemorrhagic fever in both humans and primates. However, the precise transmission dynamics between the natural reservoir and humans or primates are insufficiently understood, though exposure to contaminated excretions from fruit bats is likely a contributing factor (6). The virus is transmitted from human to human, particularly through direct contact with body fluids like blood, saliva, semen, and other body fluids from infected individuals (7).

The Marburg viral genome consists of 19,114 nucleotides encoding for seven proteins, including glycoprotein (GP), nucleoprotein (NP) virion protein 30 (VP30), VP35, VP24, VP40 and large viral polymerase (3).

Genomic characterization of MARV strains has revealed substantial genetic diversity across its global phylogeny, but exhibits limited genomic variation within outbreaks (8).

Sequencing of the Marburg genome has provided deeper insights into viral evolution and identified critical mutation hotspots during adaptation to new hosts, particularly in the VP40 protein and the NP-VP35 intergenic region (9). These mutations can influence the virus’s ability to evade the immune system, therefore enhancing its pathogenicity.

### Statement of the specific scope of the study

In this study, we used a near real-time genomics sequencing approach to characterize the Marburg virus in blood samples obtained from patients in the ongoing outbreak in Rwanda. Our aim was to use the genomic sequencing data to identify and characterize the virus and understand its evolution and transmission dynamics during the current outbreak. This research underscores the importance of continuous genomic surveillance which not only facilitates outbreak tracking but also informs public health strategies for effectively controlling this deadly pathogen.

## Results

Phylogenetic analyses support that the outbreak resulted from a single zoonotic transmission event with limited human-to-human transmission rather than multiple independent zoonotic transmission events (**Figure 1A**). Multiple independent zoonotic transmissions would have resulted in a set of more diverged sequences introduced from the genetically diverse viral population in the reservoir (10). The probable index case was a man in his 20’s with an occupational exposure to fruit bats (*Rousettus aegyptiacus*) in a mining cave environment, who exhibited symptoms highly consisting with the classic clinical presentation of MVD. Additionally, serological testing of three contacts of the index case exhibited MARV antibodies, further supporting epidemiological tracing of the index case. The outbreak lineage is most closely related to a sequence sampled in Kampala, Uganda in September 2014 from a healthcare worker (HCW), with no secondary cases observed (KP985768). The source of the zoonotic transmission in 2014 was never identified (11). However, the Rwanda-Uganda sister lineages are significantly diverged from one another, separated by 82 nucleotide substitutions (**Figure 1A**). In Bayesian phylogenetic reconstructions, we estimate that the two lineages diverged from a common ancestor that circulated in the animal reservoir in November 2008 [95% highest posterior density, or HPD: May 2007 and June 2010] (**Supplementary Figure 1 A-B**).

**Figure 1:**
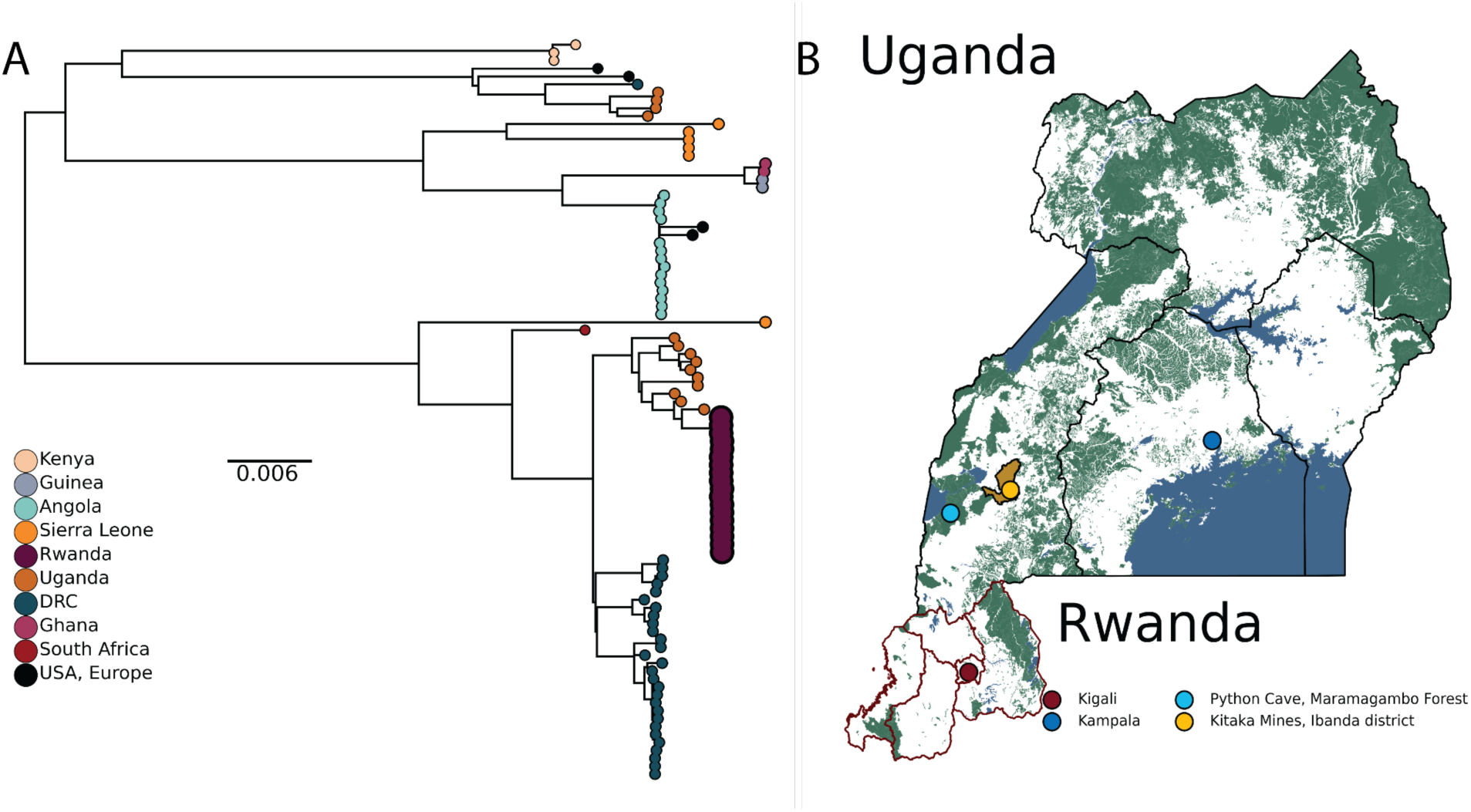
A) Maximum likelihood phylogeny of the global MARV dataset. Tips are annotated by country of isolation. This study’s tips are relatively enlarged. B) Map of Uganda and Rwanda. The city of patients in this study (Kigali) is annotated in dark red. Python Cave and Kitaka Mine in Ibanda district in southwestern Uganda are annotated in light blue and yellow, respectively.

The outbreak lineage in Rwanda is nested within a larger clade that includes diversity sampled from bats in southwestern Uganda from 2007 onwards, along with two human cases identified in Uganda in 2017 (**Figure 1A, B**). The closest bat virus (JX458855) was sampled from a juvenile Egyptian fruit bat (*Rousettus Aegyptiacus)* in 2009 from Python Cave in Queen Elizabeth National Park, a popular tourist attraction in south western Uganda (**Figure 1B**) (6). The lineage in Rwanda is significantly diverged from JX458855, sharing a common ancestor that likely circulated in the reservoir as early as June 2006 (95% HPD: December 2004 to November 2007) (**Supplementary Figure 1 A-B**). *Rousettus aegyptiacus* are a cave dwelling species, with spillover events and limited outbreaks frequently associated with mining activities. This includes outbreaks and sporadic cases linked to Python cave and the Kitaka mine that is only 50 km away in 2007 and 2008, which both hosts *Rousettus aegyptiacus* colonies of over 50 000 bats. (6,10,12). The divergent relationship of the Rwandan Marburg virus outbreak lineage indicates that the virus’s dispersal through host networks involved larger scale animal movement over a long period of time, rather than a direct ancestral connection to the *Rousettus aegyptiacus* colony in southwestern Uganda.

The index patient’s close relative was admitted to Hospital 1 (H1) at the end of August 2024 but succumbed to the illness prior to the definitive diagnosis of MVD. It is highly probable that this patient infected healthcare workers (HCWs) at H1, where MVD was subsequently confirmed at the end of September 2024. From 22-28 September, 26 cases were reported in HCWs at H1 and H2, respectively. The limited genetic variation among the outbreak sequences was consistent with a very recent common ancestor, the short sampling period, and previous observations from the Angola MARV outbreak (**Figure 2A**) (12). A cluster of eight identical sequences was sampled, spanning the full sampling period (C1 in **Figure 2A, B**). The majority of the C1 sequences were isolated from healthcare workers (HCWs) at H1 from 22-28 September, with reported symptom onset as early as mid-September (**Figure 2B**). A C1 sequence was sampled from a HCW at H2 at the end of September, suggesting infection from an unsampled C1 case in H2 or dissemination between the two healthcare facilities. There are recorded cases of healthcare workers moving between healthcare settings H1 and H2. This is further supported by cluster C2, consisting of two healthcare workers from H1 and H2 respectively that share the synonymous SNP C11070T relative to C1. Both C2 cases report symptom onset and were sampled in the later weeks of the epidemic. There are a further seven identical sequences (C4) sampled between H1 and H2 across the full sampling period that are one synonymous SNP (T10031C) away from C1. C002, a healthcare worker employed at both H1 and H2, is believed to have acquired the infection at H1. This individual subsequently became the index case at H2, leading to a nosocomial cluster at H2 and infections at H1. Overall, there were only three synonymous SNPs observed in the outbreak clade: A5856G (n=1 sequence), T10031C (n=7), and C11070T (n=2). A5856G and T10031C as well as C11070T are outside of the CDS of the GP and VP24 genes, respectively. There is therefore no evidence that post-emergence substitutions enhanced human transmission.

**Figure 2:**
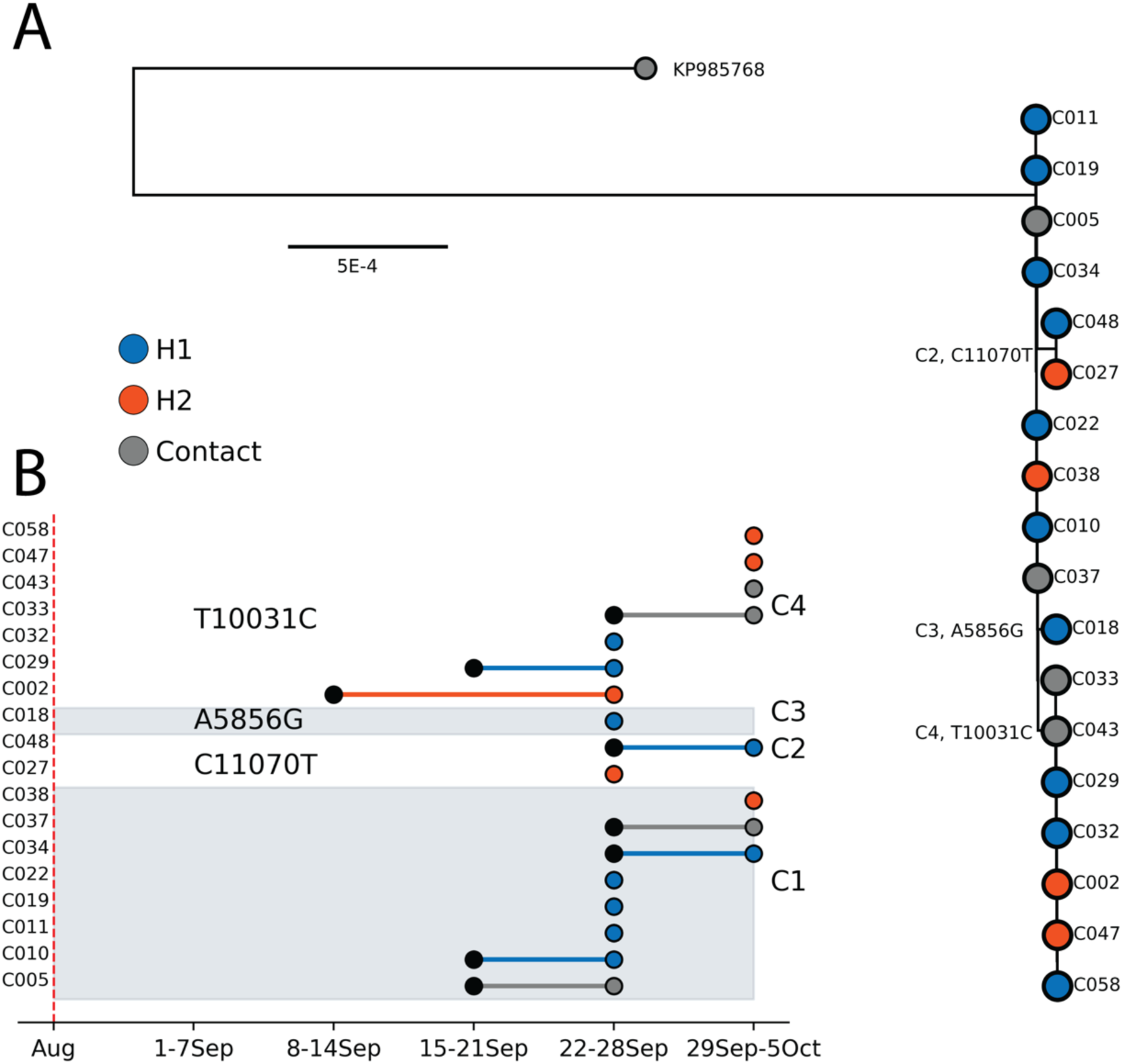
A) Maximum likelihood phylogeny of the outbreak clade, with the Kampala outgroup. Sequences are colored with associated hospital. SNPs reconstructed relative to the common ancestor are annotated in text, as are the clusters defined by the SNP presence. The coordinates are relative to NC_001608 B) Epidemiological timeline of the sampled sequences in Figure 2A, as annotated on the y-axis. The first black marker, if present, indicates the week of symptom onset if distinct from week of sampling, with the second indicating the week of sampling. Sequences are colored by their associated hospital (H1, H2), and partitioned by SNP presence into clusters, as annotated in text. Red dashed line indicates the contact of the index case admitted to H1.

An exhaustive investigation of all cases was conducted which led to identifying the index case. This in-depth process involved a comprehensive collection of diverse data sources including clinical records, travel history, antibody serology testing results, and contact tracing information. This evidence revealed that the earliest patient was traced back to a mining cave and exhibited symptoms highly consisting with the classic clinical presentation of MVD. Additionally, serology testing results from three contacts linked to the index case exhibited antibodies of persons exposed to Marburg virus. This suggests that the mining cave may have served as a potential habitat for the MARV animal reservoir, particularly the fruit bat (*Rousettus aegyptiacus*). Following up on these findings, the Rwanda Ministry of Health through a multisectoral collaboration launched an investigation surveillance with the aim of screening fruit bats, mainly cave dwellers, for Marburg virus (**Supplementary Figures 2 A, B and C**).

## Discussion

The Rwanda MVD outbreak exhibited substantial human-to-human transmission, primarily involving healthcare personnel, as is often observed with viral hemorrhagic fever (VHF) outbreaks closely associated with nosocomial and occupational infections(18). However, Rwanda effectively contained the spread from further reaching the broader community. Positive cases were promptly isolated and treated with monoclonal antibodies and antivirals, achieving a favorable case fatality rate of approximately 23%. This underscores the ongoing need to enhance healthcare personnel’s knowledge and attitudes regarding VHF case management.

Through evolutionary and phylogenetic analysis, we observed that the early outbreak sequences represented very limited genomic variation, indicating that the outbreak originated from a single zoonotic transmission event. The Rwandan lineage shared a common ancestor with sequences originating from diversity sampled in bats in southwestern Uganda, thought the lineages had diverged over decades in the animal reservoir. It is likely that enhanced zoonotic surveillance will reveal many unsampled intermediates that can clarify the ecological pathways of transmission between bat colonies in Uganda and Rwanda. Previous research indicates that bats can harbor MARV for extended periods, with active mining areas in Rwanda providing ideal habitats for bats and increasing the likelihood of zoonotic transmission (19).

We observed three synonymous single nucleotide polymorphisms (SNPs) outside the coding regions of the GP and VP24 genes, respectively. Studies have shown that the Marburg virus VP24 protein interacts with the nucleoprotein (NP) and other cellular membranes, facilitating the release of new virions from infected cells (20). Additionally, research highlights that MARV VP24 directly interacts with the human and bat Keap1 proteins, which modulate antioxidant responses to support viral replication, a strategy likely critical for viral persistence and host adaptation (21). Genomic sequencing of MARV in Rwanda has yielded critical insights into viral circulation dynamics. However, as these SNPs fall outside of coding regions, it is unlikely that they functionally contribute to enhanced viral fitness on any scale. Our results should be interpreted within the context of our smaller sample size, which only covers the first two weeks of the outbreak.

This research underscores the importance of continuous genomic surveillance which not only facilitates outbreak tracking but also informs public health strategies for effectively controlling this deadly pathogen. In addition, previous research indicates that bats can harbor MARV for extended periods, with active mining areas in Rwanda providing ideal habitats for bats and increasing the likelihood of zoonotic transmission. Furthermore, Rwanda’s location in a region of consistent turbulent outbreaks, including Ebola, Rift Valley fever, COVID-19, dengue, and Marburg, underscores the importance of regional and international collaboration to enhance outbreak preparedness and responses for global health security.

## Acknowledgments

We gratefully acknowledge the support of the Government of Rwanda and the Ministry of Health, through the Rwanda Biomedical Center, for facilitating and supporting this sequencing work. We extend our sincere appreciation to both the African Centre of Excellence for Genomics of Infectious Diseases (ACEGID) and the Africa Centre for Disease Control and Prevention (Africa CDC) for their invaluable contributions. This work was made possible through the collaborative efforts of development partners and stakeholders, to whom we are deeply grateful.

## Author contributions

Conceptualization: Y.B, L.M, S.N, C.H, C.M.M, Methodology: Y.B, L.M, A.A, E.U, R.M, E.P, P.S, C.H, C.M.M Investigation: I.M, E.R, E.K, E.R, N.G Sampling: R.S, P.G, P.S, J.C.M PCR Experiment: E.K, F.U, V.M, G.I, H.M Sequencing: Y.B, L.M, E.U, L.I, C.M, A.U, I.U, R.N, A.U, J.K, J.P.M Bioinformatics Analysis: E.P, R.M, J.P.M, N.G, J.R, A.O, C.H Lab Enablement: S.T.K, A.A, O.O, O.A, A.H, C.M.M Writing-Original Draft: Y.B, L.M, C.M, R.M, J.P.M, P.S Writing Review and Editing: Y.B, L.M, E.E.S.F, E.R, C.M, M.S, M.N, T.T, D.T, E.S, M.G, J.P.R, J.N, A.R, F.K, G.M, A.A, S.N, C.H, C.M.M.

## Diversity, equity, ethics, and inclusion [optional]

NA

## Data availability

All the sequences are available on NCBI - GenBank under Accession Number PQ552725 (https://www.ncbi.nlm.nih.gov/nuccore/PQ552725). All other data are available upon request on https://github.com/rbx-bfx/illumina_vsp.

## Materials and Methods

### Ethics declaration

The study was approved by the Rwanda National Ethics Committee (FWA Assurance No. 00001973 IRB 00001497 of IORG0001100-Protocol approval notice: N° 121/RNEC/2024).

### Patient sample collection

We obtained whole blood samples from suspected patients presented with clinical symptoms (high fever, severe headaches, muscle aches, fatigue, nausea, vomiting, and diarrhea) of MDV. Sample testing was performed at Rwanda Biomedical Centre/National Reference Laboratory whereby samples were kept in a cold chain prior to plasma separation and analysis.

### Nucleic acid extraction

Viral RNA was extracted from 140µl plasma using the QIAamp Viral RNA Mini Kit (Qiagen, Hilden, Germany), following the manufacturer’s instructions adapted to the in-house standard operating procedures with an elution volume of 60µl. The extracted RNA was quantified using a Qubit fluorometer, and the samples were stored at −80°C until further analysis.

### RT-PCR and genomic sequencing

For the detection and amplification of the target regions of the MVD, we employed the use of RealStar® Filovirus Screen RT-PCR Kit 1.0 (Altona Diagnostics, Hamburg, Germany), which is based on real-time PCR technology, for the qualitative detection and differentiation of Ebola and Marburg virus specific RNA in human EDTA plasma. The RT-qPCR was conducted following manufacturer’s user guide and performed on CFX96 BIORAD machine. The amplification followed an initial denaturation at 95°C for 5 minutes, followed by 40 cycles of denaturation at 95°C for 30 seconds, annealing at 55°C for 30 seconds, and extension at 72°C for 1 minute, concluding with a final extension at 72°C for 5 minutes. The amplified products were analyzed via 2% agarose gel electrophoresis to confirm successful amplification and appropriate product size. Following amplification, library construction was performed on 30 samples using the Illumina RNA Prep with Enrichment (L) Tagmentation (IRPE) workflow and the lllumina Viral Surveillance Panel v2 kit (VSP v2 kit). Total RNA extracts were converted to cDNA, tagmented and thereafter amplified. Genomic region of interest was captured using a hybrid capture method. Probes were isolated using magnetic pulldown which are selectively enriched for the desired regions. Enriched libraries were quantified using dsDNA HS assay and Qubit fluorometer. Libraries were thereafter denatured and normalized at a final loading concentration of 0.8pM. Paired-end sequencing was performed using a NextSeq 550 with a 300-cycle mid-output cartridge, with the sequencing depth aimed at a minimum coverage of 100x to ensure robust variant detection.

### Bioinformatics Analysis

#### Genome Assembly

We employed a reference-based genome assembly pipeline to analyze the sequencing data generated using the Illumina Viral Surveillance Panel (VSP) v2 Kit (described above). This workflow integrates several steps including; quality control, host genome filtering, and viral genome assembly to accurately reconstruct viral sequences from targeted sequencing.

To ensure that only high-quality data were processed downstream, raw sequencing reads were first processed with *fastp v. 0.23.24* (*13*) for trimming adapters and filtering out low-quality bases. Following this, human host genome sequences were filtered out by aligning the cleaned reads to the human genome (hg38) using *Bowtie2 v.2.5.4* (14). Unmapped reads, presumed to be of viral origin, were retained for downstream analysis. These de-hosted reads were then aligned to a Marburg virus reference genome (NC_001608.3) using *Minimap2 v.2.28* (*15*), The sequences had 70-97% genomic coverage. We manually masked two mutations in respective consensus sequences that were adjacent to potential misalignment.

### Variant Calling and Consensus Generation

Variant calling was performed using both *ivar 1.4.3* (*16*) and *LoFreq v.2.1.5* (*17*). A depth coverage threshold of 50 reads per nucleotide position was applied to ensure robust and reliable variant detection. This threshold was also applied to the generation of consensus sequences which was also done using *ivar 1.4.3*.,

### Phylogenetic

We combined our eighteen higher quality sequences (coverage >= 70%) with all publicly available MARV sequences available on Genbank (N=81). To root the tree, we included 8 Ravn virus sequences included as an outgroup, which were pruned from the tree and excluded in all subsequent analyses. We aligned the sequences using *Mafft v 7.52* (*22*) and reconstructed a phylogenetic tree using *IQTree 2.2.5* (*23*) under modelfinder plus (*24*), with ultrafast bootstrapping (25). We performed the initial ancestral state reconstruction using *Treetime v0.9.3* (*26*).

### Bayesian phylogenetic reconstruction

The current genomic data does not provide sufficient temporal signal to estimate the rate of evolution in the outbreak clade. However, we were not interested in estimating the time to the most recent common ancestor (tMRCA) of the outbreak clade, as determining the emergence timing of the outbreak clade will more likely depend on the epidemiological and contact history of the index case. We wanted to estimate tMRCA of the outbreak clade and its closest human and zoonotic outgroups to better understand its potential zoonotic emergence pathway. We therefore reconstructed the time-resolved phylogeny under a fixed local clock. We assumed a fixed value of 1e-3 substitutions per site per year as a plausible rate of evolution for a RNA virus in the early stage of an outbreak for the defined outbreak clade. We allowed the remainder of the tree to evolve under a lognormal prior centered on 5e-4. (27) We used a two-phase coalescent model: the tree from the MRCA (Rwandan lineage) onward was modeled with an exponential growth model, with the earlier phase modeled as a constant-population size coalescent model. We ran two independent chains of 100 million states to ensure convergence, discarding the initial 10% of each chain as burn-in. The chains were then combined with LogCombiner. For all subsequent analyses, we assessed convergence using Tracer, and constructed a maximum clade credibility (MCC) tree in TreeAnnotator 1.10 (28).

## Supplemental Figures

**Supplemental Figure 1:**
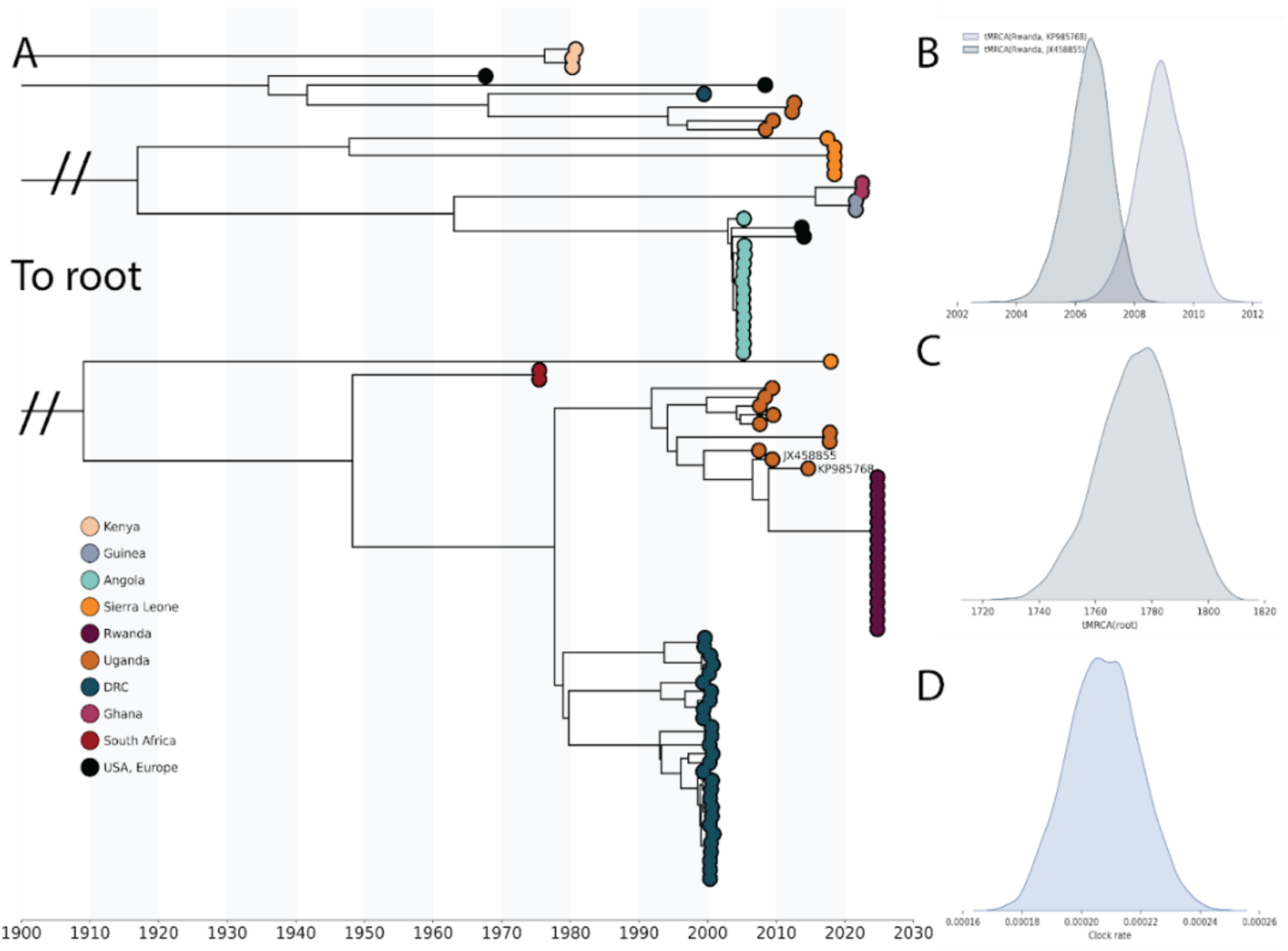
A) Time resolved maximum clade credibility tree of global MARV dataset, with closest human (KP985768) and zoonotic (JX458855) sequence to the Rwandan outbreak lineage annotated. B) tMRCA of the Rwandan lineage and KP985768 as well as JX458855 respectively. C) tMRCA of root, which is truncated in A for clarity. D) Estimate clock rate in substitutions per site per year for full tree, excluding the outbreak clade.

**Supplemental Figure 2:**
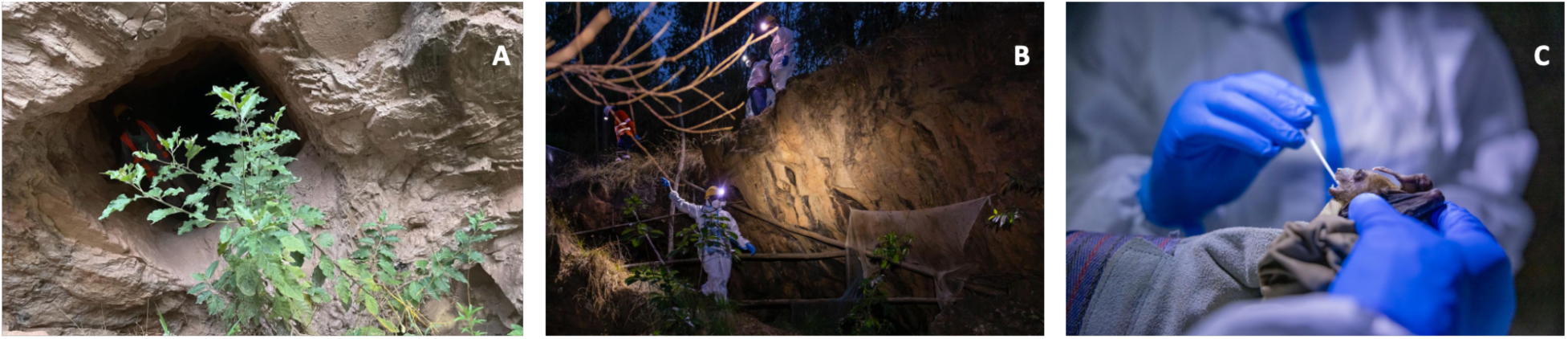
A) Mining cave entrance with human activity; B) Trapping fruit bats with net; C) Sampling bats for Marburg virus.

## Reference

1. Mehedi M, Groseth A, Feldmann H, Ebihara H. Clinical Aspects of Marburg Hemorrhagic Fever. Future Virol [Internet]. 2011 Sep [cited 2024 Oct 24];6(9):1091–106. Available from: https://www.tandfonline.com/doi/full/10.2217/fvl.11.79

2. Kilangisa LM, Max BL, Kayuni EA, Shao ER, Mashauri HL. Marburg virus disease: lesson learned from the first outbreak encounter in Tanzania. International Journal of Surgery: Global Health [Internet]. 2023 Jul [cited 2024 Oct 17];6(4). Available from: https://journals.lww.com/10.1097/GH9.0000000000000186

3. Paton, A. W., & Paton, J. C. (2018). Detection and characterization of Shiga toxigenic *Escherichia coli* by PCR. In D. Liu (Ed.), Molecular Medical Microbiology (2nd ed., pp. 213–239). Academic Press. 10.1016/B978-0-12-812026-2.00013-X

4. Srivastava S, Sharma D, Kumar S, Sharma A, Rijal R, Asija A, et al. Emergence of Marburg virus: a global perspective on fatal outbreaks and clinical challenges. Front Microbiol [Internet]. 2023 Sep 13 [cited 2024 Oct 14];14:1239079. Available from: https://www.frontiersin.org/articles/10.3389/fmicb.2023.1239079/full

5. Ministry of health R. Stop the Marburg virus [Internet]. 2024 Oct. Available from: https://x.com/rwandahealth/status/1846968616278778288?s=12&t=HFAKR78RPRo-Cmz7j7ciDQ

6. Amman BR, Carroll SA, Reed ZD, Sealy TK, Balinandi S, Swanepoel R, et al. Seasonal Pulses of Marburg Virus Circulation in Juvenile Rousettus aegyptiacus Bats Coincide with Periods of Increased Risk of Human Infection. Kawaoka Y, editor. PLoS Pathog [Internet]. 2012 Oct 4 [cited 2024 Oct 18];8(10):e1002877. Available from: https://dx.plos.org/10.1371/journal.ppat.1002877

7. Amman BR, Schuh AJ, Albariño CG, Towner JS. Marburg Virus Persistence on Fruit as a Plausible Route of Bat to Primate Filovirus Transmission. Viruses [Internet]. 2021 Nov 30 [cited 2024 Oct 18];13(12):2394. Available from: https://www.mdpi.com/1999-4915/13/12/2394

8. Towner JS, Khristova ML, Sealy TK, Vincent MJ, Erickson BR, Bawiec DA, et al. Marburgvirus Genomics and Association with a Large Hemorrhagic Fever Outbreak in Angola. J Virol [Internet]. 2006 Jul [cited 2024 Oct 19];80(13):6497–516. Available from: https://journals.asm.org/doi/10.1128/JVI.00069-06

9. Wei H, Audet J, Wong G, He S, Huang X, Cutts T, et al. Deep-sequencing of Marburg virus genome during sequential mouse passaging and cell-culture adaptation reveals extensive changes over time. Sci Rep [Internet]. 2017 Jun 13 [cited 2024 Oct 19];7(1):3390. Available from: https://www.nature.com/articles/s41598-017-03318-3

10. Towner JS, Amman BR, Sealy TK, Carroll SAR, Comer JA, Kemp A, et al. Isolation of Genetically Diverse Marburg Viruses from Egyptian Fruit Bats. Fouchier RAM, editor. PLoS Pathog [Internet]. 2009 Jul 31 [cited 2024 Oct 25];5(7):e1000536. Available from: https://dx.plos.org/10.1371/journal.ppat.1000536

11. Nyakarahuka L, Ojwang J, Tumusiime A, Balinandi S, Whitmer S, Kyazze S, Kasozi S, Wetaka M, Makumbi I, Dahlke M, Borchert J, Lutwama J, Ströher U, Rollin PE, Nichol ST, Shoemaker TR. Isolated Case of Marburg Virus Disease, Kampala, Uganda, 2014. Emerg Infect Dis. 2017 Jun;23(6):1001-1004. doi: 10.3201/eid2306.170047. PMID: 28518032; PMCID: PMC5443453

12. Carroll SA, Towner JS, Sealy TK, McMullan LK, Khristova ML, Burt FJ, et al. Molecular Evolution of Viruses of the Family Filoviridae Based on 97 Whole-Genome Sequences. J Virol [Internet]. 2013 Mar [cited 2024 Oct 25];87(5):2608–16. Available from: https://journals.asm.org/doi/10.1128/JVI.03118-12

13. Chen S. Ultrafast one-pass FASTQ data preprocessing, quality control, and deduplication using fastp. iMeta [Internet]. 2023 May [cited 2024 Oct 24];2(2):e107. Available from: https://onlinelibrary.wiley.com/doi/10.1002/imt2.107

14. Langmead B, Salzberg SL. Fast gapped-read alignment with Bowtie 2. Nat Methods [Internet]. 2012 Apr [cited 2024 Oct 25];9(4):357–9. Available from: https://www.nature.com/articles/nmeth.1923

15. Li H. Minimap2: pairwise alignment for nucleotide sequences. Birol I, editor. Bioinformatics [Internet]. 2018 Sep 15 [cited 2024 Oct 25];34(18):3094–100. Available from: https://academic.oup.com/bioinformatics/article/34/18/3094/4994778

16. Grubaugh ND, Gangavarapu K, Quick J, Matteson NL, De Jesus JG, Main BJ, et al. An amplicon-based sequencing framework for accurately measuring intrahost virus diversity using PrimalSeq and iVar. Genome Biol [Internet]. 2019 Dec [cited 2024 Oct 25];20(1):8. Available from: https://genomebiology.biomedcentral.com/articles/10.1186/s13059-018-1618-7

17. Wilm A, Aw PPK, Bertrand D, Yeo GHT, Ong SH, Wong CH, et al. LoFreq: a sequence-quality aware, ultra-sensitive variant caller for uncovering cell-population heterogeneity from high-throughput sequencing datasets. Nucleic Acids Research [Internet]. 2012 Dec 1 [cited 2024 Oct 25];40(22):11189–201. Available from: https://academic.oup.com/nar/article/40/22/11189/1152727

18. Raab, M., Pfadenhauer, L.M., Millimouno, T.J., et al. Knowledge, attitudes and practices towards viral haemorrhagic fevers amongst healthcare workers in urban and rural public healthcare facilities in the N’zérékoré prefecture, Guinea: a cross-sectional study. BMC Public Health 20, 296 (2020). 10.1186/s12889-020-8433-2

19. Scarpa F, Bazzani L, Giovanetti M, Ciccozzi A, Benedetti F, Zella D, Sanna D, Casu M, Borsetti A, Cella E, et al. Update on the Phylodynamic and Genetic Variability of Marburg Virus. Viruses. 2023; 15(8):1721. 10.3390/v15081721

20. Abir, M. H., Rahman, T., Das, A., Etu, S. N., Nafiz, I. H., Rakib, A.,…Hassan, M. M. (2022). Pathogenicity and virulence of Marburg virus. Virulence, 13(1), 609–633. 10.1080/21505594.2022.2054760

21. Edwards MR, Johnson B, Mire CE, Xu W, Shabman RS, Speller LN, Leung DW, Geisbert TW, Amarasinghe GK, Basler CF. The Marburg virus VP24 protein interacts with Keap1 to activate the cytoprotective antioxidant response pathway. Cell Rep. 2014 Mar 27;6(6):1017–1025. doi: 10.1016/j.celrep.2014.01.043. Epub 2014 Mar 13. PMID: 24630991; PMCID: PMC3985291.

22. Kazutaka Katoh, Daron M. Standley, MAFFT Multiple Sequence Alignment Software Version 7: Improvements in Performance and Usability, Molecular Biology and Evolution, Volume 30, Issue 4, April 2013, Pages 772–780, 10.1093/molbev/mst010

23. Bui Quang Minh, Heiko A Schmidt, Olga Chernomor, Dominik Schrempf, Michael D Woodhams, Arndt von Haeseler, Robert Lanfear, IQ-TREE 2: New Models and Efficient Methods for Phylogenetic Inference in the Genomic Era, Molecular Biology and Evolution, Volume 37, Issue 5, May 2020, Pages 1530–1534, 10.1093/molbev/msaa015.

24. Kalyaanamoorthy, S., Minh, B., Wong, T. et al. ModelFinder: fast model selection for accurate phylogenetic estimates. Nat. Methods 14, 587–589 (2017). 10.1038/nmeth.4285.

25. Diep Thi Hoang, Olga Chernomor, Arndt von Haeseler, Bui Quang Minh, Le Sy Vinh, UFBoot2: Improving the Ultrafast Bootstrap Approximation, Molecular Biology and Evolution, Volume 35, Issue 2, February 2018, Pages 518–522, 10.1093/molbev/msx281.

26. Pavel Sagulenko, Vadim Puller, Richard A Neher, TreeTime: Maximum-likelihood phylodynamic analysis, Virus Evolution, Volume 4, Issue 1, January 2018, vex042, 10.1093/ve/vex042.

27. Carroll SA, Towner JS, Sealy TK, McMullan LK, Khristova ML, Burt FJ, Swanepoel R, Rollin PE, Nichol ST. Molecular evolution of viruses of the family Filoviridae based on 97 whole-genome sequences. J Virol. 2013;87:2608–2616. doi: 10.1128/JVI.03118-12.

28. Bouckaert, R., Heled, J., Kühnert, D., Vaughan, T., Wu, C-H., Xie, D., Suchard, MA., Rambaut, A., & Drummond, A. J. (2014). BEAST 2: A Software Platform for Bayesian Evolutionary Analysis. PLoS Computational Biology, 10(4), e1003537. doi:10.1371/journal.pcbi.1003537.

