## Supplemental_Figure_1 for "Genomic characterization uncovers transmission dynamics of Marburg Virus in Rwanda following a single zoonotic spillover event": Supplemental_Figure_1_MARV_Genomic_Characterization_Rwanda.pdf

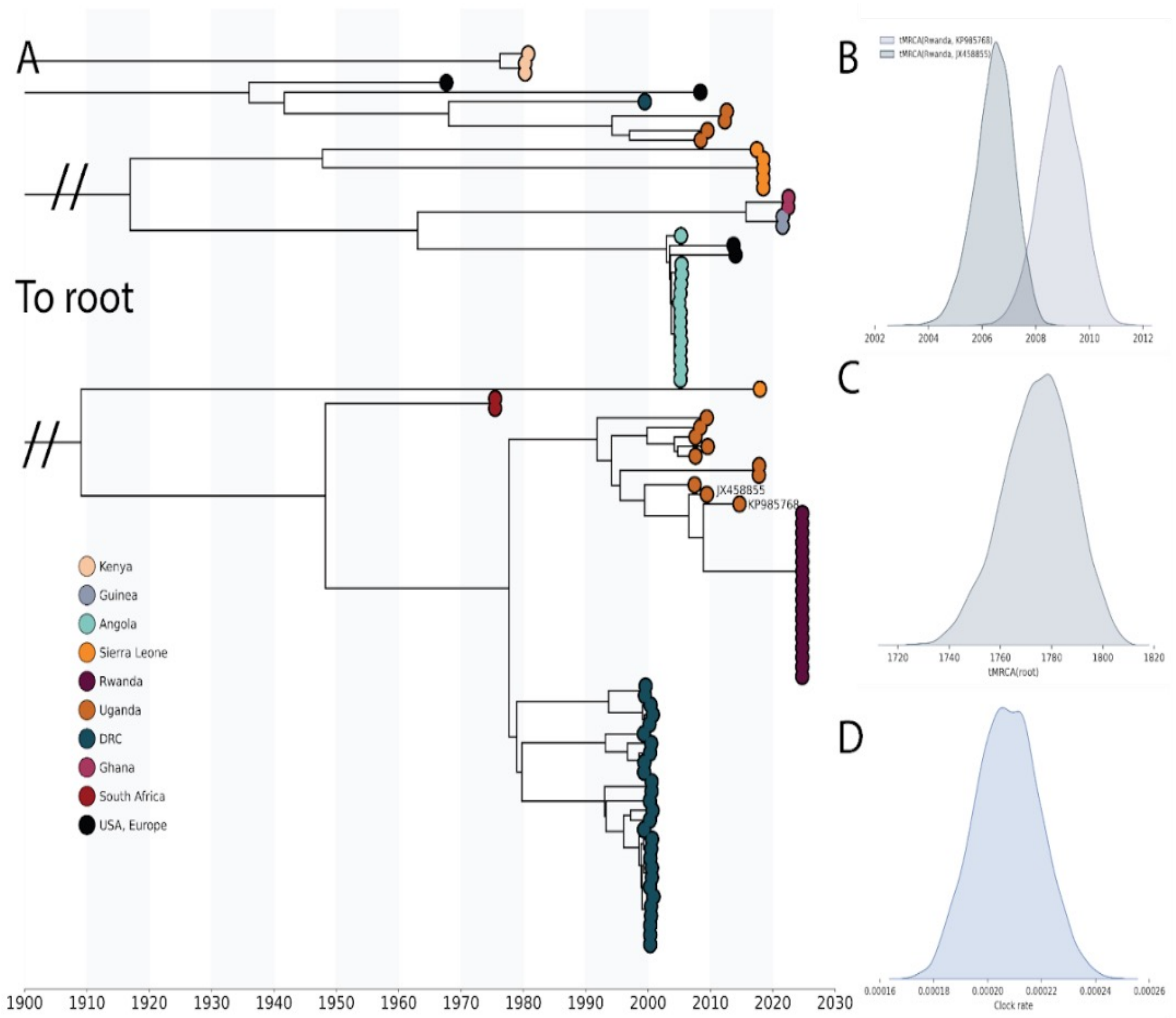


**Supplementary Figure 1:** *A) Time resolved maximum clade credibility tree of global MARV dataset, with closest human (KP985768) and zoonotic (JX458855) sequence to the Rwandan outbreak lineage annotated. B) tMRCA of the Rwandan lineage and KP985768 as well as JX458855 respectively. C) tMRCA of root, which is truncated in A for clarity. D) Estimate clock rate in substitutions per site per year for full tree, excluding the outbreak clade.*
