## Supplemental_Figure_2 for "Genomic characterization uncovers transmission dynamics of Marburg Virus in Rwanda following a single zoonotic spillover event": Supplemental_Figure_2_MARV_Genomic_Characterization_Rwanda.pdf

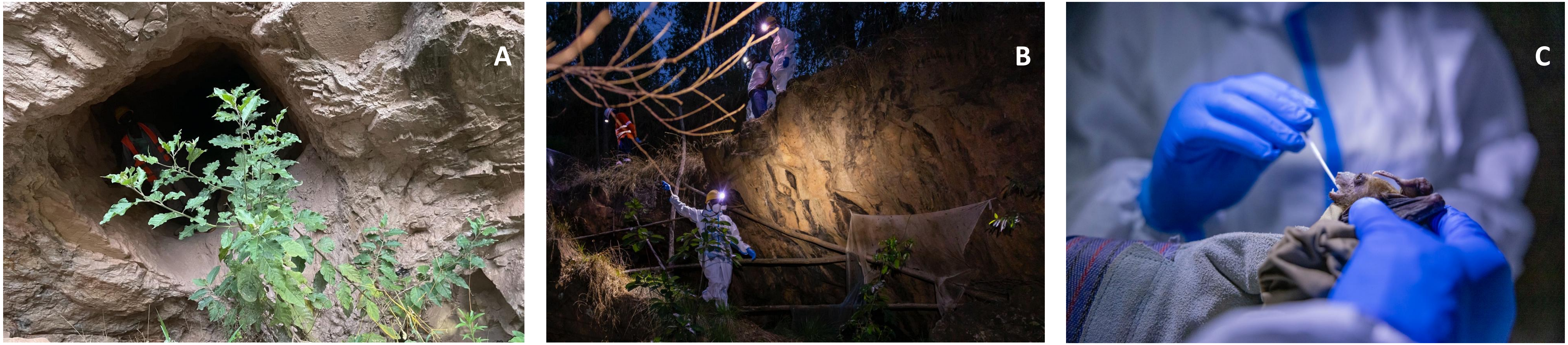


**Supplemental Figure 2:** *A) Mining cave entrance with human activity; B) Trapping fruit bats with net; C) Sampling bats for Marburg virus.*
